# External validation of self-supervised transfer learning for noninvasive molecular subtyping of pediatric low-grade glioma using T2-weighted MRI

**DOI:** 10.64898/2026.01.27.26344883

**Authors:** Jay J. Yoo, Divyanshu Tak, Khashayar Namdar, Matthias W. Wagner, Anthony Liu, Uri Tabori, Cynthia Hawkins, Birgit B. Ertl-Wagner, Benjamin H. Kann, Farzad Khalvati

**Author notes:** Jay J. Yoo and Divyanshu Tak are co-first authors. Benjamin H. Kann and Farzad Khalvati are co-senior authors.

## Abstract

**Purpose:** To externally evaluate three binary classification models designed to differentiate the molecular subtype of pediatric low-grade glioma (pLGG) between BRAF Fusion, BRAF Mutation, and Wild Type on T2-weighted magnetic resonance imaging using self-supervised transfer learning, which enables effective performance in a low data setting.

**Materials and methods:** This retrospective study evaluates pLGG molecular subtyping models, pre-trained using data collected at Dana Farber Cancer Institute/Boston’s Children’s Hospital, on two datasets from the Hospital for Sick Children, one consisting of patients identified from the electronic health record between January 2000 to December 2018 (n=336) and another consisting of patients identified from the electronic health record between January 2019 to April 2023 (n=87). These datasets consist of T2-weighted MRI with pLGG and corresponding genetic marker identifications, labelled as BRAF Fusion, BRAF Mutation, or Wild Type. The datasets included manually annotated ground-truth segmentations that were used in the classification pipeline during evaluation. The models were evaluated using the area under the receiver operating characteristic curve (AUC). To acquire a per-class probabilities across all three considered molecular subtypes, we used the output probabilities from each binary model as logits input to a Softmax function. These probabilities were used to determine the AUC of the models on each evaluated dataset.

**Results:** The models performed achieved a macro-average AUC of 0.7671 on the newer dataset from the Hospital for Sick Children but achieved a lower macro-average AUC of 0.6463 on the older dataset from the Hospital for Sick Children.

**Conclusions:** The evaluated pLGG molecular subtyping models have the potential for effective generalization but may require further fine-tuning for consistent performance across varying datasets.

## 1 Introduction

Pediatric low-grade gliomas (pLGG) are the most commonly occurring brain tumors among children, with pLGGs comprising approximately 40% of central nervous system tumors in children [1]. Modern targeted therapeutic strategies for treating pLGGs require accurate identifications of the molecular status of the pLGGs [2, 3]. The two most common genetic alterations that occur in pLGG are KIAA1549-BRAF fusion (BRAF Fusion) and BRAF p.V600E (BRAF Mutation). Recent advances in BRAF-directed therapies have placed increased importance on both the identification of BRAF subtypes and the differentiation between BRAF altered and non-BRAF altered (Wild Type) genetic markers [4, 5].

The molecular subtype of pLGG can be determined using biopsy and surgical resection [2, 6], but these approaches can be infeasible or inadvisable. Biopsy and surgical resection can be costly, have associated risks such as neurological compromise, leave residual tumors, and can fail due to insufficient sample acquisition [3, 6]. These risks incentivize the development and use of imaging-based non-invasive solutions.

Tak et al. from the Kann Laboratory within the Artificial Intelligence in Medicine Program (AIM) at Dana Farber Cancer Institute, Mass General Brigham, Harvard Medical School, proposed a training approach that combines self-supervision and transfer learning to improve the training of classification models that identify the BRAF status from MRI in low data settings [5]. Their proposed deep learning pipeline first preprocesses T2-weighted MRI and then inputs the resultant preprocessed, skull-stripped images to a nnUNet-based 3D tumor segmentation model [7]. Three ResNet-based binary molecular subtype classifiers (BRAF Fusion vs. rest; BRAF Mutation vs. rest; Wild Type vs. rest) are used to generate predictions on the likelihood BRAF Fusion, BRAF Mutation, or a Wild Type pLGG is present in the image. The predictions are aggregated using a consensus decision block to output the molecular subtype prediction and its probability. The binary classifiers were trained using an approach referred to as TransferX. TransferX begins with pretrained weights from RadImageNet [8] but then applies two additional stages of finetuning on separate but relevant classification tasks. The classification tasks that these two finetuning stages are trained serve as pretext tasks for self-supervision. Once these finetuning stages are completed, a final finetuning step using the target class is applied [5].

In this work, we evaluated the performance of the molecular subtyping classification models from Tak et al., which we will refer to as the pLGG molecular subtyping models, on external datasets of T2-weighted MRI capturing pLGG without any additional training. This collaborative work further validates the molecular subtype classification models from Tak et al. on T2-weighted MRI of pLGG from the Hospital for Sick Children (SickKids).

## 2 Materials and methods

### 2.1 Datasets and preprocessing

Two datasets from SickKids were used to externally validate the pLGG molecular subtyping models. This study conformed to the guidelines and regulations of the research ethics board of The Hospital for Sick Children, which approved this retrospective study and waived the need for informed consent due to the retrospective nature of the study. All data used in this study was de-identified after acquisition from the electronic health record database.

The first dataset consists of 336 T2-weighted MRI 3D images containing pLGG. These images were acquired from patients ranging from 0 to 18 years of age who were identified using the electronic health record database of SickKids from January 2000 to December 2018. These images are labeled with the pLGG genetic markers, which were simplified to BRAF Fusion, BRAF Mutation, and Wild Type. This dataset will be referred to as SK1.

The second dataset consists of 87 T2-weighted MRI 3D images containing pLGG from patients ranging from 0 to 18 years of age. These patient images were identified using the electronic health record database of SickKids from January 2019 to April 2023. As was done with the first dataset, these images were also labeled with the pLGG genetic markers, simplified to BRAF Fusion, BRAF Mutation, and Wild Type. This dataset will be referred to as SK2.

The two datasets had ground-truth tumor segmentations available, which were used instead of whereas the proposed approach from Tak et al. used outputs from a nnUNet-based 3D tumor segmentation model. For the purposes of this study, the available ground-truth tumor segmentations were used instead of predictions from a 3D tumor segmentation model.

The two datasets were preprocessed by first resampling the images to 1 × 1 × 1*mm*^3^ voxel size using linear interpolation before applying rigid registration using a pediatric MRI template sourced from the National Institutes of Health MRI Study of Normal Brain Development (NIHPD) Objective 1 Atlases (29) as was done by Boyd et al. [7]. Skull stripping was performed after the registration and N4 Bias field correction was applied once the images were skull stripped. The registration was done using the SimpleITK Python package, the skull stripping was done using the HD-BET Python package, and the bias field correction was done using SimpleITK.

### 2.2 Evaluation details

For this study, we evaluated the pLGG molecular subtyping models using the area under the receiver operating characteristic curve (AUC). When computing the AUC, the output probabilities from each model were considered as logits, and then the Softmax function was applied to these logits to achieve per-class probability predictions. The per-class probability predictions were then used to acquire the corresponding AUCs. The macro-average AUC across the three per-class AUCs was also evaluated. In addition to the AUC metrics, the classification accuracy was acquired by assigning the predicted class as the class with the greatest probability after applying the Softmax function and comparing the prediction to the ground truth. A decision threshold of 0.5 is used to predict the accuracy which results with lower accuracies compared to when an optimized decision threshold is used.

## 3 Results

Table 1 presents the performance of the pLGG molecular subtyping models across the evaluated datasets. The pLGG molecular subtyping models demonstrated strong performance on the SK2 dataset but presented decreased performance on the SK1 dataset.

**Table 1:** AUCs and accuracies of the pLGG molecular subtyping models across the SK1 and SK2 datasets.

| Dataset | AUC |  |  |  | Accuracy |
| --- | --- | --- | --- | --- | --- |
|  | BRAF Fusion | BRAF Mutation | Wild Type | Macro-average |  |
| SK1 | 0.6892 | 0.6580 | 0.5917 | 0.6463 | 0.4464 |
| SK2 | 0.7423 | 0.8209 | 0.7381 | 0.7671 | 0.6092 |

## 4 Discussion

This study demonstrated that the evaluated pLGG molecular subtyping models are capable of effective generalization without additional training as demonstrated by their performance on the SK2 dataset. However, the results on the SK1 dataset show limitations in the capability of the pLGG molecular subtyping models to classify all MRI. Variations between datasets such as institutional data drift can negatively impact the performance of the models. Therefore, the models would benefit from fine-tuning before use in external sites. The original study from Tak et al. presented the ability for the deep learning pipeline they proposed to effectively train the model in low data settings [5]. The deep learning pipeline could therefore be adapted to fine-tune the models on small local datasets prior to deployment at external sites.

Future work will expand on the external validation of the pLGG molecular subtyping models, and investigate the impact of fine-tuning on the performance of the pLGG molecular subtyping models when applied to external datasets. In addition, the Kann Laboratory proposed a nnUNet-based 3D tumor segmentation model that was used as part of the molecular subtyping pipeline [7]. Once exploration of the molecular subtype classification models is complete, the segmentation models will be externally evaluated and validated.

## Data Availability

The datasets analysed in the present study are not publicly available due to institutional policy.

## Notes

### Competing Interest Statement

The authors have declared no competing interest.

### Funding Statement

This Study was supported by the Chair in Medical Imaging and Artificial Intelligence, a joint Hospital-University Chair between the University of Toronto, The Hospital for Sick Children, and the SickKids Foundation.

### Author Declarations

This study conformed to the guidelines and regulations of the research ethics board of The Hospital for Sick Children, which approved this retrospective study and waived the need for informed consent due to the retrospective nature of the study.

